# Hepatitis A virus whole genome sequencing strategy using NGS/Illumina technology

**DOI:** 10.1101/2024.05.05.24306642

**Authors:** Paola Sicilia, Anabella C. Fantilli, Facundo Cuba, Guadalupe Di Cola, María Gabriela Barbás, Tomás Poklepovich, Viviana E. Ré, Gonzalo Castro, María Belén Pisano

**Affiliations:** Departamento Laboratorio Central de la Provincia de Córdoba, Ministerio de Salud, Gobierno de la Provincia de Córdoba. Tránsito Cáceres de Allende 421, X5000HVE, Córdoba, Argentina; Instituto de Virología “Dr. J. M. Vanella”, CONICET, Facultad de Ciencias Médicas, Universidad Nacional de Córdoba. Enfermera Gordillo Gómez s/n, Ciudad Universitaria, CP5016, Córdoba, Argentina; Unidad Operativa Centro Nacional de Genómica y Bioinformática - ANLIS “Dr. Carlos G. Malbrán”. Av. Vélez Sarsfield 563, C1282 AFF, Ciudad Autónoma de Buenos Aires (CABA), Argentina

**Keywords:** hepatitis A virus, HAV, NGS, sequencing, whole genome sequencing

## Abstract

**Background:** Molecular epidemiology of hepatitis A virus (HAV) plays a critical role in identifying outbreak origin and conducting surveillance. Although it is mostly carried out using short partial VP1/2A genomic sequences, utilizing whole-genome sequences (WGS) provides more accurate and robust information.

**Objective:** we aimed to develop an amplicon-based next-generation sequencing (NGS) strategy to obtain complete HAV genomes utilizing the COVIDSeq Test (Illumina), available in surveillance laboratories after the COVID19 pandemic.

**Study design:** 25 primer pairs were designed and used to amplify partial genomic fragments (400bp) that comprise the entire HAV genome sequence from HAV previously positive serum and stool samples from Argentina. The DNA library was prepared using the Illumina COVIDSeq Test and sequenced in a MiSeq equipment. Phylogenetic analyses were performed with IQ-Tree using WGS and VP1/2A partial sequences of 1084pb and 422pb.

**Results:** 11 samples could be successfully amplified and sequenced, with horizontal coverage between 79.3%-100% (>90% in 9 samples). Individual RT-PCRs and Sanger sequencing with specific primers had to be performed in 6 samples to cover gaps of Ns. Phylogenetic analyses of WGS and VP1/2A partial sequences yielded similar results (clustering with genotype IA), although 422pb fragments showed low accurate grouping definition.

**Conclusion:** the amplicon-based NGS whole genome sequencing tool developed by adapting the COVIDSeq test to HAV, proved to be efficient in generating new complete and near-complete viral sequences. The study of the 1084bp fragment of the VP1/2A region would constitute a useful alternative option for outbreak investigation with public health impact.

## Introduction

Hepatitis A (HA) is an acute inflammation of the liver caused by the hepatitis A virus (HAV), a member of the *Picornaviridae* family and the *Hepatovirus* genus (Lee et al., 2023). The virus is transmitted by the fecal-oral route, mainly by contaminated food and water, as well as through person-to-person contact (World Health Organization (WHO), 2023). HAV is a non-enveloped virus with a single stranded 7.5 kb genome that contains 5’ and 3’ noncoding regions, one open reading frame (ORF), and a poly(A) tract (Batista et al., 2020; Lee et al., 2022). Genetic variability has led to the identification of seven HAV genotypes (I-VII), with genotypes I to III and VII infecting humans, including seven sub-genotypes (IA, IB, IC, IIA, IIB, IIIA, and IIIB) (Lee et al., 2022; Vaughan et al., 2014).

Molecular epidemiology of HAV plays a critical role in the epidemiologic surveillance, viral source tracking, the identification of outbreaks, understanding the introduction of genetically diverse strains from geographically distant regions and genotype displacements (Lee et al., 2022; Vaughan et al., 2014). In this context, phylogenetic and genotypic analyses of HAV have traditionally been performed based on partial-genome sequences, including the VP1/P2A junction, VP1/P2B junction, C termini of VP3, N terminus of VP1, entire VP1, and 5’ UTR regions(Aguirre et al., 2011; Jansen et al., 1990; Lee et al., 2022; Nainan et al., 2005; Robertson et al., 1991; Yanez et al., 2014; Yilmaz et al., 2017). Analysis of this sub-genomic regions has been proven to be useful for outbreak investigations and surveillance. However, comparing short sequences among epidemiologically unrelated strains suggests that specific identification of HAV strains for molecular surveillance can be achieved only using whole-genome sequences (WGS), mainly through enhanced resolution phylogenetic trees, which helps to identify closely related viruses within transmission networks (Cleary et al., 2023; Vaughan et al., 2014). Additionally, whole-genome sequences can offer insights into punctual mutations, their combinations, and recombination events (Aguirre et al., 2011).

Next generation sequencing (NGS) has emerged as a robust platform for identifying and characterizing pathogens, to investigate population demographics, etiologic agents and outbreak locations, allowing for the acquisition of more comprehensive information than Sanger sequencing in a relatively short period of time, including the generation of complete genomes (Lee et al., 2023). This was notably demonstrated during the COVID19 pandemic, when these techniques were widely employed to study SARS-CoV-2 variants. However, to date, there are few protocols using this technology to rapidly and efficiently generate HAV whole-genome sequences for genotyping and molecular epidemiological tracking (Cleary et al., 2023; Zufan et al., 2024).

In Argentina, as well as in other South American countries, a shift in HAV endemicity has been registered due to socioeconomic and sanitary improvements, together with the introduction of official vaccination programs (Andani et al., 2020). The implementation of the immunization scheme in our country started in 2005, consisting of a single-dose in 12-month-old children. Consequently, the notification of clinical cases of HA has drastically decreased (Ministry of Health of Argentina, 2019), resulting in lower incidence rates, sporadic cases, and limited outbreaks in unvaccinated individuals over 20 years of age (Andani et al., 2020). However, more recently, clinical cases have been recorded among young adults aged 20 to 39 years old, in whom a greater susceptibility to HAV infection has been reported (Andani et al., 2020; Ministry of Health of Argentina, 2019; Yanez et al., 2014). In this context, from mid-2016 to early 2018, several hepatitis A outbreaks disproportionately affected unvaccinated young adult men, particularly men who have sex with men (MSM), in Europe, the USA and South America, including central Argentina (Mariojoules et al., 2019). This is a clear example on how improvements in sanitary conditions and vaccine introduction can alter the viral circulation pattern and its ecological niche. It is known that the immunological pressure generated by vaccines can induce genetic changes or the emergence of new variants. Additionally, migration and the globalization of travel favors the introduction of new variants from other regions of the world, as evidenced by the recent introduction of HAV strains from Europe to America in specific population groups (Mariojoules et al., 2019).

In this context, the analysis of complete viral genomes offers insights into the identification of genetic variants that may be associated with changes in virulence, transmissibility and vaccine escape. Moreover, it facilitates decision-making in public health by enhancing the understanding of outbreaks, viral evolution and key aspects of epidemiological surveillance. In Argentina, where HAV has been under vaccine pressure since 2005, local sequence data are scarce, with only one complete Argentinean HAV genome available to date (Aguirre et al., 2011). Furthermore, there are few published whole genome sequencing protocols worldwide that use NGS technology. With this background, we aimed to develop a strategy for HAV whole-genome sequencing by adapting the NGS Illumina COVIDSeq test, available in many surveillance laboratories, and to obtain HAV whole genome sequences from the central region of Argentina.

## Materials and methods

### Samples

To implement the methodology, we selected 16 previously obtained plasma (n=11) and stool (n=5) samples positive for HAV by RT-real time PCR (Pintó et al., 2009). Some of them had also been amplified by RT-Nested PCR and sequenced (Fantilli et al., 2023; Mariojoules et al., 2019). Before analysis, stool samples were clarified with PBS at a 10% dilution and centrifuged at 10,000 rpm for 30 min, taking the supernatant for further analysis.

### Molecular methods and NGS sequencing

An amplicon deep sequencing strategy was carried out using the adapted Illumina COVIDSeq Test (USA). Firstly, nucleic acid extraction was carried out from 200 µL of each sample with the High Pure Viral Nucleic Acid kit (Roche), following the manufacturer’s instructions. The extracted RNA was then annealed in a step of 65°C for 3 min using random hexamers to prepare for cDNA synthesis. cDNA was then synthesized using the First Strand cDNA Master Mix provided in the COVIDSeq kit under the following conditions: 5 min at 25°C, 10 min at 50°C and 5 min at 80°C.

To perform sample enrichment, 25 primer pairs were designed to amplify partial genomic fragments of approximately 400 bp that comprise the entire HAV genome sequence, using the Primal Scheme program (https://primalscheme.com/) with the sequences under the GenBank accession numbers X83302 (HAV genotype IA reference sequence) and HM769724 (complete HAV genome from Argentina) as reference genomes (Table 1). The primers were then evaluated *in silico* using the MEGA v11 (Tamura et al., 2021a) and OligoAnalyzer (Integrated DNA Technologies, https://www.idtdna.com/calc/analyzer) programs. Based on the nucleotide differences between the 2 HAV complete genomes used for the design, 9 primers were degenerated (at the divergent positions, Table 1). Afterwards, each pair of primers was subjected to PCRs individually to check for individual amplification. Since the protocol generates a series of overlapping PCR amplicons, multiplex PCRs were carried out using 4 pools of primers at a concentration of 10-13 µM (Supplementary Material 1), interspersing the even primers with the odd ones (so that they do not hybridize between them) (pools 1A and 1B: odd primers; pools 2A and 2B: even primers). Four PCRs were then performed using each pool of primers and the Illumina PCR Mix provided in the kit, under the following conditions: 98°C for 3 min, followed by 35 cycles of 98°C for 15 sec and 63°C for 5 min.

**Table 1.**
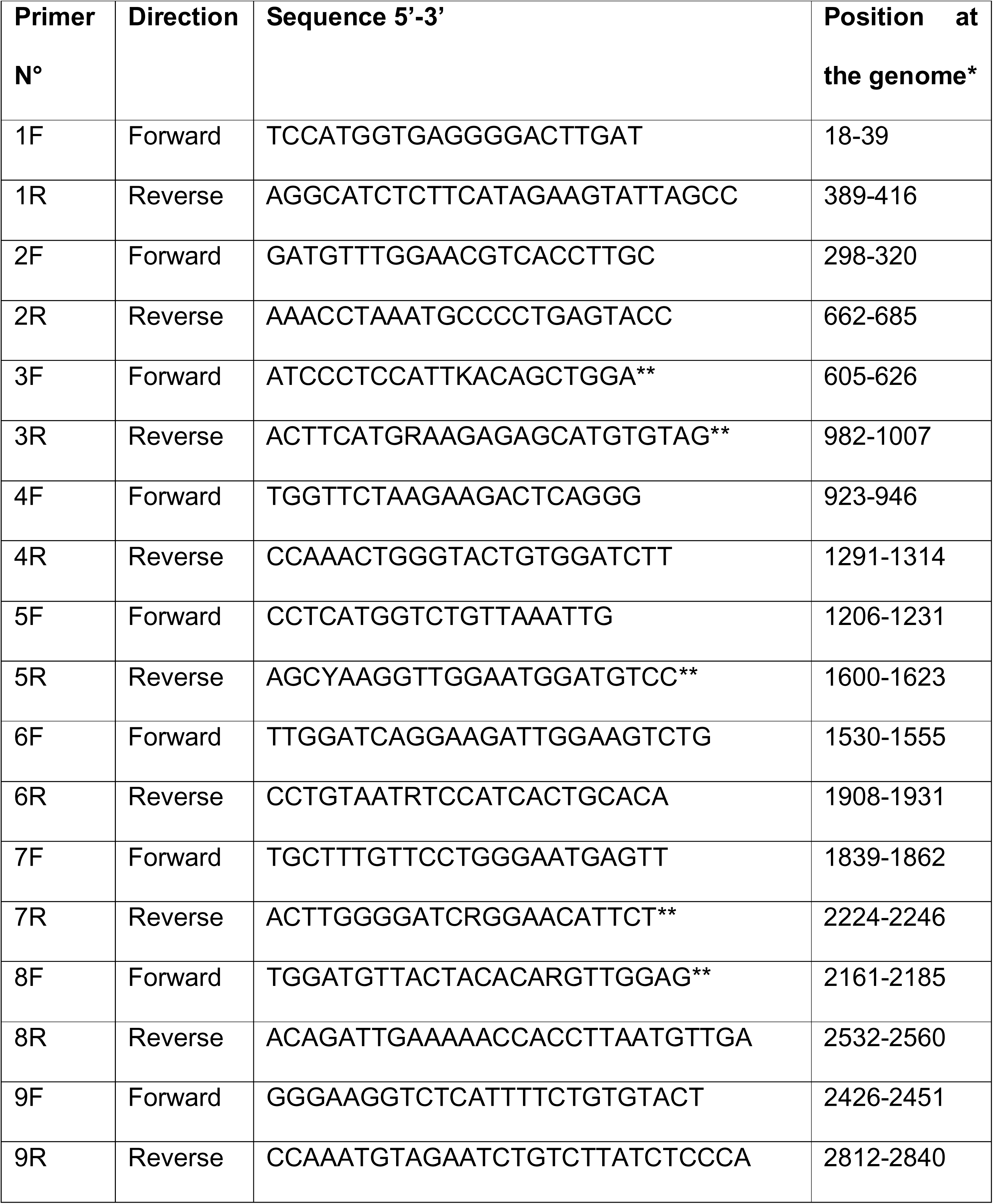
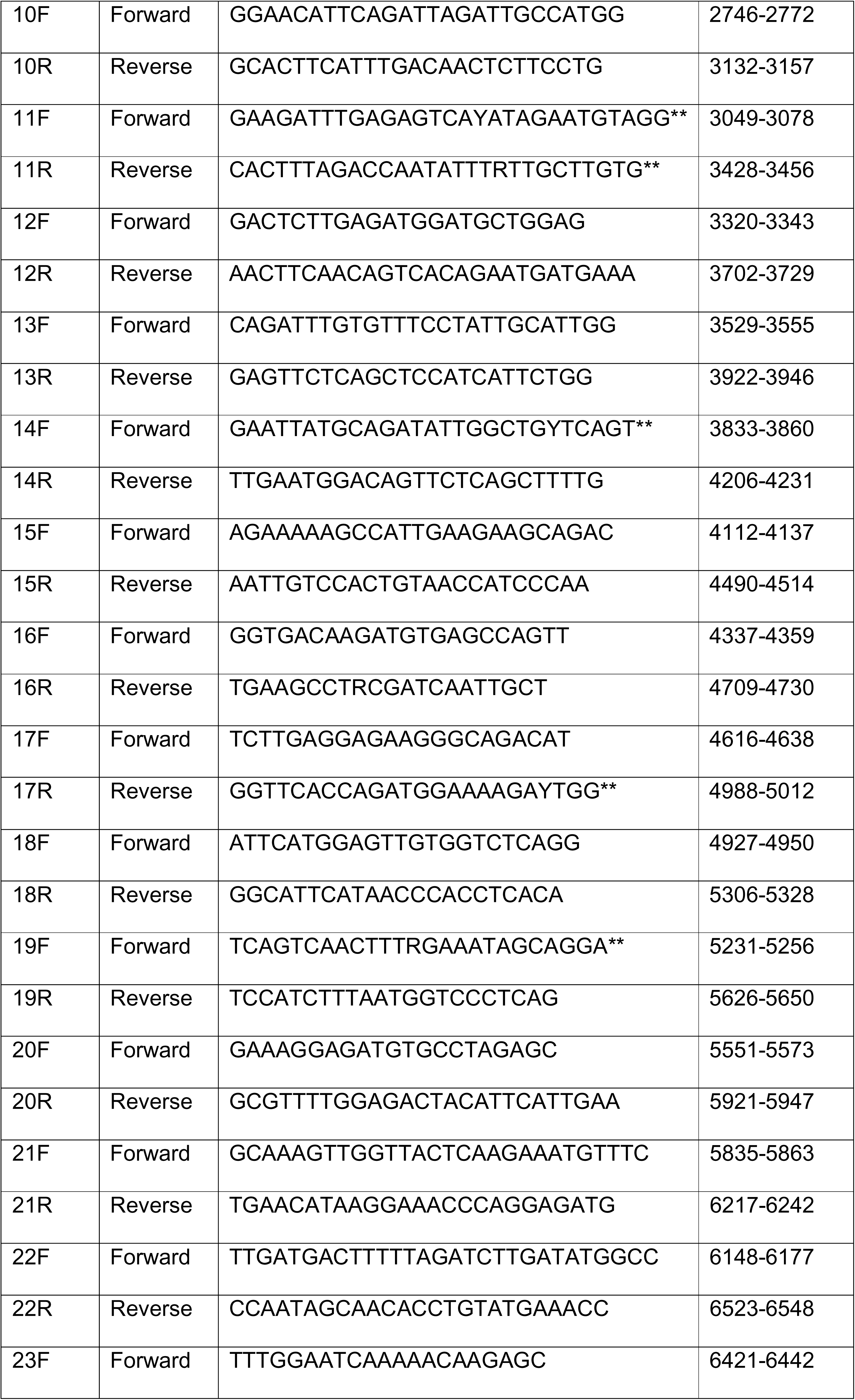
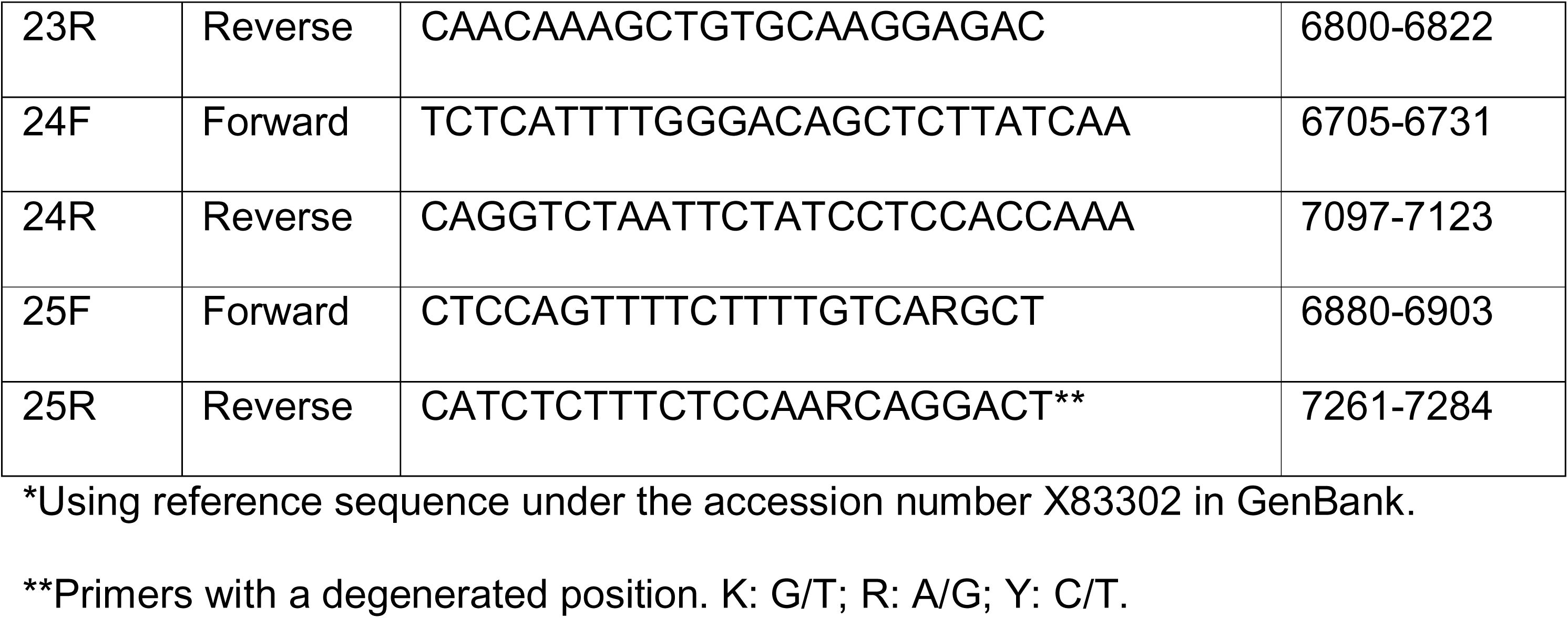
Primers designed and used for HAV amplification before NGS sequencing.

The DNA library was prepared using the Illumina COVIDSeq Test (USA), and sequencing was conducted on a MiSeq equipment. Quality control of the sequencing data was performed using FastQC (Andrews S., 2010) and Trim Galore (Krueger, 2007). The curated paired-end FASTQ files were aligned to the reference genome (Accession N°: X83302), and consensus sequences were generated using BWA (Li, 2013), SAMtools (Danecek et al., 2021), and iVar (Grubaugh et al., 2019) tools. A minimum depth of 10X was required, and positions with lower depth were represented by the nucleotide ‘N’.

### Sanger sequencing

After NGS sequencing, some of the samples presented gaps in the generated sequence data (generated in some positions by <10X depth). To fill these gaps, conventional RT-PCRs with the corresponding specific designed primers from this study were carried out, followed by Sanger sequencing. Each RT-PCR was performed using the LunaScript® RT SuperMix kit (New England BioLabs2, USA) for retrotranscription and the Platinum PCR Super Mix High Fidelity (Invitrogen, USA) for the PCR, under the following conditions: a first denaturation of 94°C for 2 min, 35 cycles of 94°C for 15 sec, 55°C for 30 sec, 68°C for 1 min and 50 sec adding 5 sec per cycle, and a final extension of 68°C for 7 min. PCR products were then purified with the PureLink Quick Gel Extraction kit (Invitrogen, USA) and sequenced in an ABI automatic sequencer (3500xL Genetic Analyzer - Applied Biosystems).

Whole or near-whole genome sequences obtained were uploaded to the NCBI GenBank database under the accession numbers PP661694 to PP661699 and PP693071 to PP693075.

### Phylogenetic analysis

The analysis included 11 HAV whole and near-whole genome sequences along with reference sequences for each HAV genotype and subtype. Three distinct trees were generated: one with the whole genome sequences (WGS) and the others with sequences from the partial VP1/2A genomic regions of 1084 bp and 415 bp. Sequences were edited and aligned with MEGA v.11 (Tamura et al., 2021b). Phylogenetic analyses were performed using the maximum-likelihood method with the software IQ-TREE v2. 1 (Minh et al., 2020; Nguyen et al., 2015); employing the best-fit model of nucleotide substitution as selected by ModelFinder (Kalyaanamoorthy et al., 2017). In this case, the analyses were performed under the TIM2+F+I+G4 (422bp and 1080bp-tree) and TIM2+F+G4 (WGS-tree) nucleotide substitution model chosen according to the Bayesian information criterion (BIC). The robustness of the phylogenetic grouping was evaluated by the SH-like approximate likelihood ratio test (SH-aLRT) (1,000 replicates) (Guindon et al., 2010) and the ultrafast bootstrap (UFB) approximation (10,000 replicates) (Hoang et al., 2018), estimated with ModelFinder in IQ-TREE. A clade with an SH-aLRT ≥ 80% and UFboot ≥ 95% was considered well-supported.

### Ethics approval

The study was conducted according to the guidelines of the Declaration of Helsinki (1964, amended most recently in 2008) of the World Medical Association, and in accordance with specific local ethical regulations (Legislature of the Province of Córdoba, 2009). The Research Evaluation Committee of the Institute of Virology “Dr. José María Vanella” (National University of Córdoba) evaluated the ethics of this study and approved its conduct. Sample IDs were not known to anyone outside the research group.

## Results

Out of the 16 samples subjected to the PCR enrichment and NGS, 11 could be successfully amplified and sequenced, all of which originally had Ct values ≤33.2 in the real time RT-PCR (Table 2).

**Table 2.**
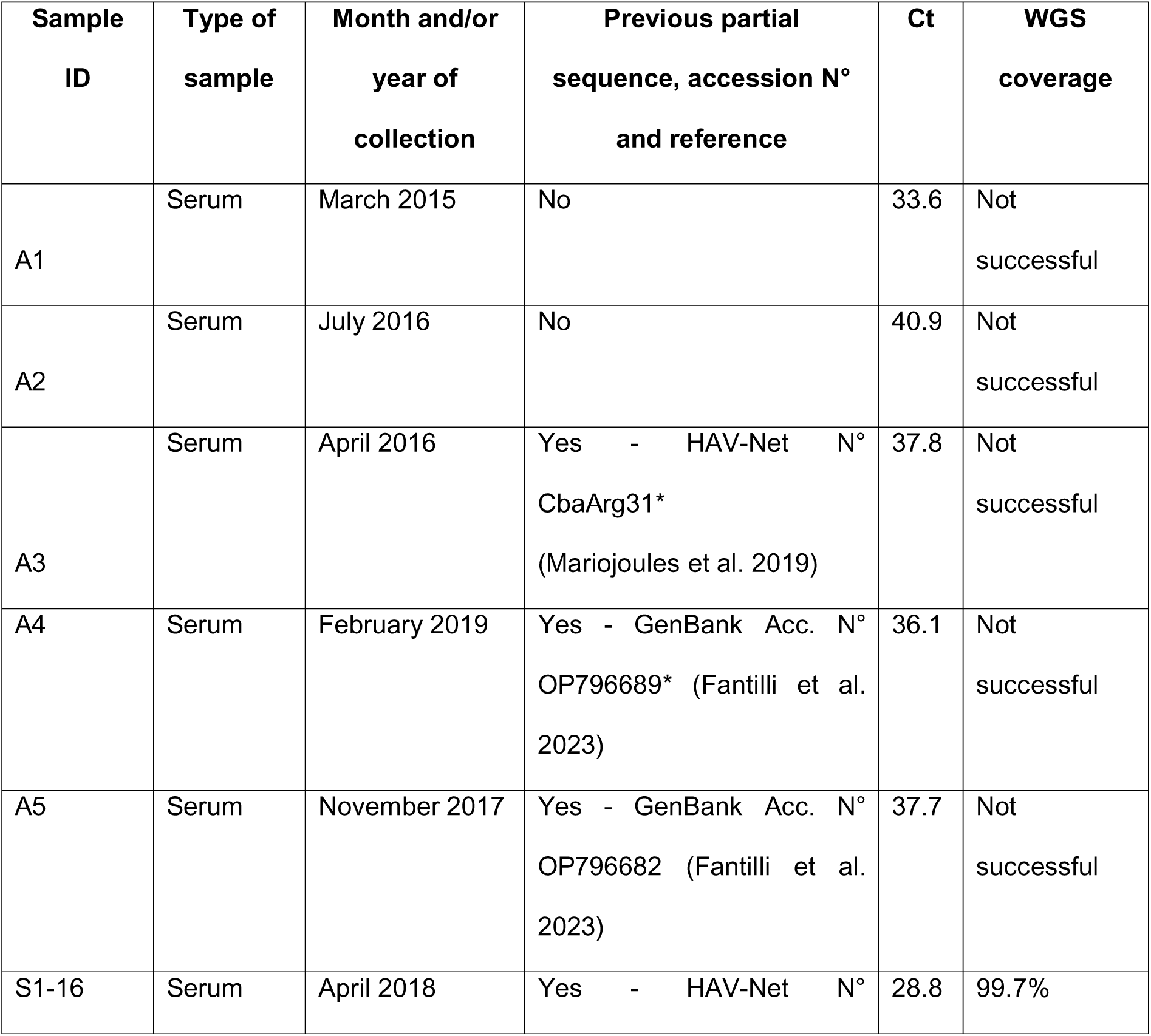
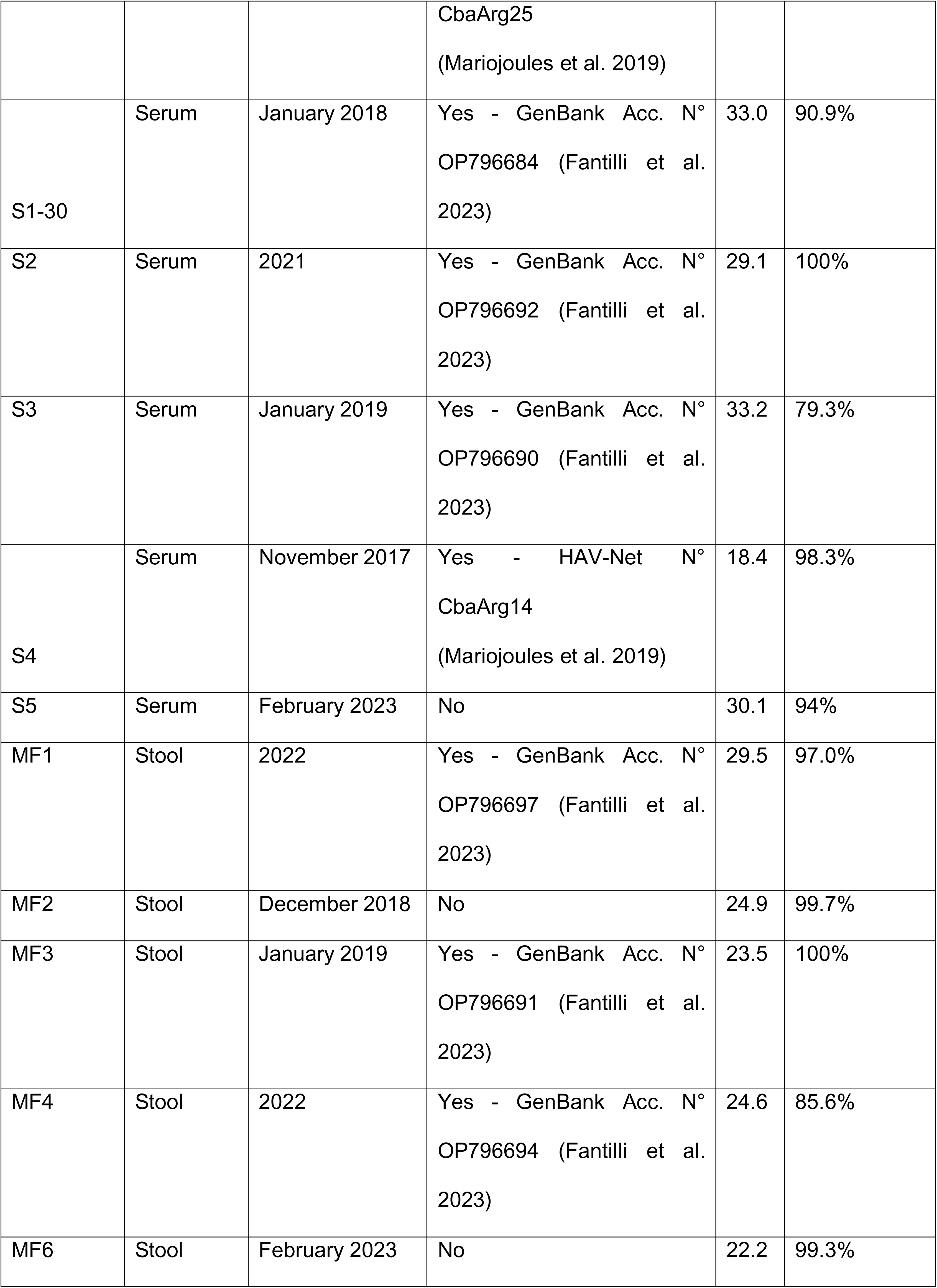
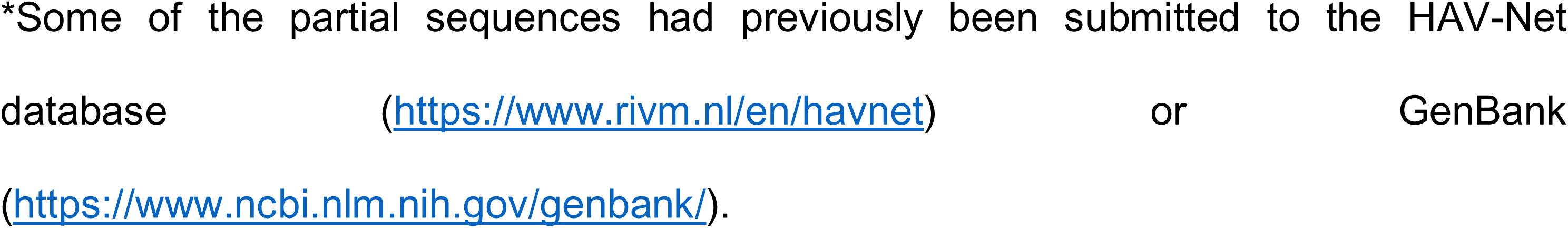
Samples included in the study and results of NGS sequencing.

NGS sequencing yielded an average of 1,509,242 reads per sample, with an estimated depth of coverage across the entire HAV genome ranging from 21,040X to 50,314X (median 41,676X). Horizontal coverage was between 90%-100% of the entire genome in 9 samples, while 2 samples exhibited lower coverage: 85.6% (sample MF4) and 79.3% (sample S3) (Table 2).

For 6 samples, individual RT-PCRs had to be performed with specific primers 4, 5, 11, 13, 14, 19 and/or 23 to cover gaps represented by Ns (regions with <10X coverage). Unfortunately, original material was unavailable for 4 of these samples to perform the assays, so they were only carried out on 2 of the specimens (Supplementary Material 2).

Figure 1 shows phylogenetic trees obtained from the analysis of both whole-genome and partial sequences from this study. Phylogenetic assessments conducted with WGS confirmed that all samples belonged to genotype IA. Sample S2 was closely related to the previous HAV Argentinean whole genome sequence, clustering within the same group alongside MF3 and S1-16. The remaining sequences clustered together, corresponding to samples isolated during the MSM outbreak of 2017-2018 and subsequent years. Analysis of the partial sequences of 422 bp and 1084 bp yielded consistent genotype classification with WGS results, albeit with lower SH-aLRT and UFBoot support values (less robustness of clades), particularly evident in the 422 bp analysis, as it was expected. However, upon conducting a more detailed analysis of the phylogenetic relationships among the isolated strains, notable differences in grouping and reduced phylogenetic support of clades were evident in the tree generated with the 422 bp sequences. In contrast, the tree generated from the 1084 bp sequences closely resembled the clustering and topology observed in the WGS analysis. Moreover, most of their branches were assigned with higher SH-aLRT and UFBoot support values, rendering these trees (WGS and 1084 bp sequences) comparable (Figure 1).

**Figure 1.**
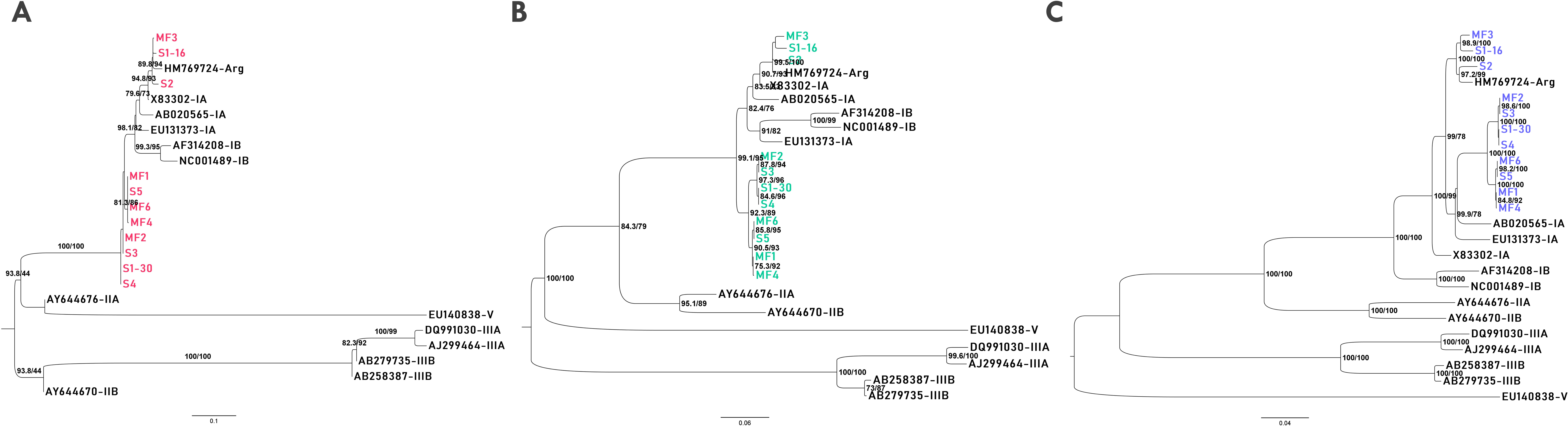
Mid-point rooted-maximum likelihood phylogenetic trees based on sequences obtained during this study (labeled in colors) and reference sequences for each genotype (black-colored tips), using 422 bp (A) and 1084 bp (B) sequences from VP1/2A and whole or near whole genome sequences (C). Statistical support [SH-aLRT (%)/UFB (%)] values are indicated at branches and nodes, and only supports over 70/70 are shown.

## Discussion

The HAV epidemiological scenario has changed worldwide after the improvement in socio-sanitary conditions as well as the introduction of the vaccine into the official vaccination schedules of numerous countries (Andani et al., 2020; World Health Organization (WHO), 2023). Thus, the epidemiology of hepatitis A has transitioned from being an endemic disease, with most of the infections and symptomatic cases during childhood, to causing sporadic outbreaks affecting young adult populations (Fantilli et al., 2023; World Health Organization (WHO), 2023). Consequently, in recent years, there have been reports of foodborne outbreaks, as well as outbreaks and sustained transmission among unvaccinated vulnerable groups, including individuals experiencing homelessness, those who inject drugs, and MSM (Zufan et al., 2024). The combination of infections in older adults and those with comorbidities has led to increase hospitalizations and deaths related to HAV infection, underscoring its emergence as a global public health concern (Mariojoules et al., 2019; World Health Organization (WHO), 2023; Zufan et al., 2024).

Molecular epidemiology has significantly contributed to understanding HAV infection sources, viral circulation, transmission routes, pathogenesis and prevention. Together with epidemiological data, it has demonstrated to improve outbreak characterization and response (Lee et al., 2022; Zufan et al., 2024). However, molecular characterization is not always performed, and, when it occurs, it typically involves partial amplification of small genomic fragments of HAV genome, such as a 450 bp genomic fragment corresponding to the VP1/2A region (Bardón De Tena et al., 2023; Freidl et al., 2017; Harries et al., 2014; Krumbholz et al., 2023; Levican et al., 2023; Mariojoules et al., 2019). Whole genome sequencing of HAV has been reported to produce a better phylogenetic signal for outbreak studies, facilitating public health interventions (Lee et al., 2022; Zufan et al., 2024). However, obtaining WGS is laborious and time-consuming when using the traditional Sanger sequencing technique. On the other hand, NGS methods have emerged as valuable tools for rapidly and efficiently generating complete genome sequences (Batista et al., 2020; Lee et al., 2023; Zufan et al., 2024). Particularly, their relevance was highlighted during the COVID-19 pandemic, when several specialized laboratories had access to this technology enabling the rapid sequencing of complete SARS-CoV-2 genomes for variant surveillance (Hare et al., 2023; Pisano et al., 2022). Despite its costliness, the widespread adoption of NGS during the pandemic paved the way for equipment acquisition, reagent availability, and professional training, thereby enabling its application for sequencing other viral genomes.

In this study, we developed a HAV whole genome sequencing protocol by adapting a SARS-CoV-2 sequencing kit (COVIDSeq test) with Illumina technology. Specific primers were designed for sample enrichment prior to sequencing, resulting in the successful acquisition of 11 HAV whole or near-whole genome sequences from clinical samples (serum and stool) from Argentina in less than 48hs, thus contributing valuable information to the only one previous complete genome available to date in our country (Aguirre et al., 2011).

Argentina, after 18 years since the introduction of the vaccine in 1-year-old children in 2005, is now considered a region with low HAV endemicity (Andani et al., 2020). Nevertheless, mirroring global trends, there has been a notable shift in the age distribution of incidence of cases in recent years, with most of them currently occurring among individuals aged 20-36 years (Fantilli et al., 2023; Zufan et al., 2024). Thus, between 2017 and 2018, outbreaks of hepatitis A were documented among MSM both in Argentina and worldwide, highlighting virus circulation despite vaccination efforts (Mariojoules et al., 2019). Furthermore, HAV has been consistently detected in local wastewater samples from 2017 to 2022, except in 2020, coinciding with the COVID-19 pandemic and associated quarantine measures and mobility restrictions (Fantilli et al., 2023). This evolving epidemiological scenario underscores the importance of continuous HAV surveillance to promptly identify outbreaks and take appropriate measures, and monitor isolated cases, focusing on genomic surveillance to detect potential mutations and emerging viral variants with increased pathogenicity or transmissibility (Fantilli et al., 2023; Zufan et al., 2024). In this context, the developed assay in this study has proven to be a useful tool for generating HAV whole or near-whole genomes, contributing to the expansion of the limited global database of HAV whole genome sequences.

A few whole genome sequencing protocols have been previously developed for HAV using NGS: three of them using primer schemes with Illumina (Lee et al., 2022), Ion Torrent (Cleary et al., 2023) and nanopore (Lee et al., 2023) technologies, another using a direct nanopore strategy (Batista et al., 2020) and a hybridization capture panel for undertaking HAV WGS using Illumina (Zufan et al., 2024). The strategy developed during this work offers an additional approach, with the advantage of using a kit for SARS-CoV-2 sequencing (and now the similar IMAP Illumina kit), commonly available in viral surveillance laboratories, thereby facilitating the acquisition of HAV sequences. Additionally, during optimization, amplification and sequencing were achievable in samples with Ct values up to 33.2, demonstrating consistent and slightly improved performance compared to some previously developed NGS-based techniques, which exhibited lower performance in samples with Ct values >30 (Cleary et al., 2023; Lee et al., 2022).

Some genomic regions could not be correctly amplified during the multiplex reaction, leading to incomplete genomes, although these gaps were successfully addressed through Sanger sequencing. However, despite these inherent challenges, the assay managed to generate whole genomes for almost half of the samples tested, achieving at least 97% of coverage in 7 samples.

In spite of phylogenetic analyses with whole genome sequences yielding similar results to those obtained from analyzing both fragments of the partial VP1/2A region, the phylogenetic signal derived from WGS was notably stronger and more robust, providing enhanced resolution and increased precision in the results (e.g.: in genotype assignment). Indeed, it has been reported that variation across the entire viral genome can significantly enhance spatial and temporal phylogenetic resolution in HAV molecular epidemiology (Cleary et al., 2023).

The phylogenetic tree generated from the analysis of partial 422 bp sequences exhibited weakly supported subtype assignments and a lack of a defined pattern and clear grouping among Argentine sequences within genotype IA. In contrast, analysis of complete genomes of these samples offered a clearer delineation of groupings, facilitating the study of phylogenetic and evolutionary relationships of the strains and, therefore, providing more evidence and information for outbreak investigation and intervention efforts. Interestingly, and unlike what happened with the short VP1/2A fragment, the phylogenetic analysis obtained using 1084 bp partial sequences produced results that were highly similar, if not identical, to those generated using WGS. This finding is of great importance for those laboratories or situations where NGS technology or resources for sequencing complete genomes are not available, but amplification and partial sequencing of the samples can still be conducted. The study of this larger fragment of the VP1/2A region would constitute a useful alternative option for outbreak investigation with public health impact.

In conclusion, the amplicon-based NGS whole genome sequencing tool, developed by adapting the COVIDSeq test to HAV, proved to be efficient in generating new complete and near-complete viral sequences from Argentina, utilizing the infrastructure, reagents and facilities acquired over the past 4 years. This approach facilitated providing solid information on the molecular epidemiology of HAV in the region. Moreover, the technique could be transferred to and used for public health surveillance, enabling focused responses to outbreaks, and potentially extending the study of alternative sample sources, such as environmental matrices.

## Supporting information

Primer preparation

Table primers for Sanger sequencing

## Data Availability

All data produced in the present study are available upon reasonable request to the authors.

