## Supplementary material for "Hepatitis A virus whole genome sequencing strategy using NGS/Illumina technology": Primer preparation

**HAV whole genome sequencing – Preparation of the primers**

Volume of stock of primers: 200µL

Primer concentration in the pool: 10µM

Primers **2, 3, 8 and 18**: faint bands, so higher primer concentration was used: 13µM

Pool of primers final volume: **200µL**

[Each primer]: 10µM 🡪 Make a dilution of 1/20 in the pool.

[Each primer]: 13µM 🡪 Make a dilution of 1/15 in the pool (primers 2, 3, 8 and 18).

**Pool 1A**: 7 primers (1, 3, 5, 7, 9, 11, 13)

- Primers 1, 5-13: 10µL of each (F y R) = 120µL

Total volume of primers = 147µL

- Primer 3: 13.5µL (F y R) = 27µL
- Water volume = 53µL
- Final volume = 200µL

**Pool 1B**: 6 primers (15, 17, 19, 21, 23, 25)

- Primers 15-25: 10µL of each (F y R) = 120µL
- Water volume = 80µL
- Final volume = 200µL

**Pool 2A**: 6 primers (2, 4, 6, 8, 10, 12)

- Primers 2 and 8: 13.5µL of each (F y R) = 54µL

Total volume of primers = 134µL

- Primers 4, 6, 10 and 12: 10µL of each (F y R) = 80µL
- Water volume = 66µL
- Final volume = 200µL

**Pool 2B**: 6 primers (14, 16, 18, 20, 22, 24)

- Primer 18: 13.5µL (F y R) = 27µL

Total volume of primers = 127µL

- Primers 14, 16, 20, 22, 24: 10µL of each (F y R) = 100µL
- Water volume = 73µL
- Final volume = 200µL
