## Supplementary material for "Hepatitis A virus whole genome sequencing strategy using NGS/Illumina technology": Table primers for Sanger sequencing

**Supplementary Material 2.** Table showing samples with regions of <10X coverage (gaps of Ns) and primer pairs used in individual RT-PCRs to complete them.

| **Sample ID** | **% coverage** | **N° primers used to fill “N”s gaps** | **Amplification** |
| --- | --- | --- | --- |
| MF1 | 97.0% | 5 | No original material. |
| MF2 | 99.7% | 4 | Yes |
| MF4 | 85.6% | 4, 5, 19, 23 | Scarce original material. Amplification with primer pair 4 and 19. |
| S1-30 | 90.9% | 4, 5, 11 | No original material. |
| S3 | 79.3% | 4, 5, 11, 13, 14, 19 | No original material. |
| S5 | 94.0% | 4, 5 | No original material. |
